# Investigating the Efficacy of Ergothioneine to Delay Cognitive Decline in Mild Cognitively Impaired Subjects: A Pilot Study

**DOI:** 10.1101/2024.07.08.24310085

**Authors:** Yu Fung Yau, Irwin K. Cheah, Rathi Mahendran, Richard M.Y. Tang, Ru Yuan Chua, Rachel E.S. Goh, Lei Feng, Jialiang Li, Ee Heok Kua, Christopher Chen, Barry Halliwell

## Abstract

**Background and objective:** Dementia, particularly Alzheimer’s disease, is a major healthcare challenge in ageing societies. Therefore, this study aimed to investigate the efficacy and safety of a dietary compound, ergothioneine, in delaying cognitive decline in elderly individuals.

**Design, intervention and measurements:** Nineteen subjects aged 60 or above with mild cognitive impairment were recruited for this double-blinded, randomized, and placebo-controlled study. Subjects received either ergothioneine (25mg per capsule) or a placebo, taken 3 times a week for one year. The whole blood profile, markers of renal and liver functions, neurocognitive performance, plasma levels of ergothioneine and its metabolites, and plasma biomarkers related to neurodegeneration were measured across the study.

**Result:** Ergothioneine intake did not alter clinical safety markers (blood counts, kidney and liver function) throughout the study, further validating its safety for human consumption. Subjects receiving ergothioneine demonstrated improved performance in assessment of learning ability and stabilized plasma levels of neurofilament light chain, compared with placebo group which saw no improvement in cognitive assessments and a significant increase in neurofilament light chain.

**Conclusion:** Prolonged intake of ergothioneine showed no toxicity in elderly individuals. Enhanced Rey Auditory Verbal Learning Test performance and stabilized neurofilament light chain levels suggest improvements in memory and learning abilities, alongside a deceleration of neuronal damage. Our results add to existing data that ergothioneine is safe for extended consumption and may hold the potential to delay cognitive decline in the elderly.

## Introduction

Dementia, especially Alzheimer’s disease (AD), presents an escalating challenge to ageing societies worldwide. The World Health Organization reported that more than 55 million people are living with dementia in 2019 and projects this number to almost triple to 153 million within the next three decades[1]. The global economic costs associated with dementia are also projected to rise from US$1.3 trillion in 2019 to US$2.8 trillion by 2030[1], necessitating more effective interventions to delay or prevent its onset.

The pathophysiology of AD is characterized by progressive neuronal loss, memory impairment, and decline in cognitive functions. Key pathological events include the accumulation of toxic amyloid oligomers and neurofibrillary tangles that lead to synaptic dysfunction and neuronal death[2–5]. Mitochondrial dysfunction, oxidative damage, inflammation, and metabolic dysfunction in neurons are key pathological mechanisms underlying the neurodegenerative cascade[6–9]. There have recently been significant advances in anti-amyloid antibody therapies, such as Lecanemab (Leqembi). Nonetheless, while this effectively clears β-amyloid in early AD patients, it only demonstrates a modest impact on disease progression and poses a significant risk of developing brain swelling and/or cerebral microhaemorrhage[10]. This underscores the pressing need to explore novel mechanisms to prevent or delay onset of dementia.

Ergothioneine (ET), a thione derivative of histidine, has recently garnered great interest for its diverse physiological properties, including antioxidant, anti-inflammatory, metal-chelating activities, and neuroprotective capabilities[11–14]. While not produced in the human body, ET can be readily absorbed from the diet, by means of a specific membrane transporter, organic transporter novel type-1 (OCTN1), which is widely expressed in the intestinal tract, and many tissues throughout the body, including the brain[15, 16], and facilitates accumulation in these tissues[17]. Studies have shown that ET can mitigate β-amyloid toxicity in neuronal cultures, *Caenorhabditis elegans* and rodent models of AD [18–22]. Moreover, we have demonstrated that individuals with mild cognitive impairment (MCI) and dementia exhibit significantly lower plasma levels of ET compared to age-matched healthy controls[23, 24]. Even more strikingly, our observational study revealed that lower plasma levels of ET in cognitively normal subjects correlated with faster cognitive decline and brain pathology on follow-up for up to 5 years[25].

Despite evidence suggesting its potential benefits in ageing and dementia from previous animal and cell studies, the direct impact of ET supplementation on cognitive health remains unexplored in humans. ET is approved for human consumption by the USA FDA and European Union (generally recognised as safe status; GRAS) as well as the European Food Safety Authority[26, 27], but has never been given for prolonged periods to elderly subjects. To bridge the gap, this pilot study aimed to explore the safety and possible cognitive benefits of ET consumption in elderly individuals with MCI. We recruited elderly participants aged between 60 - 90 years of age, from two study cohorts diagnosed with MCI and provided them with ET or placebo in a double-blinded and randomised manner. Throughout the course of the study, we examined the neurocognitive performance of the MCI subjects and their biochemical markers, including assessments of potential toxicity. We hope that our preliminary findings will inspire future research directions and larger clinical studies to evaluate the beneficial effects of ET in slowing or preventing cognitive impairment.

## Methods

### Chemicals, reagents, and investigational compound

ET, L-ergothioneine-d9 (ET-d9), L-hercynine, L-hercynine-d9, S-methyl ET were provided by Tetrahedron (www.tetrahedron.fr). Encapsulated ET (25mg/capsule Ergoneine^®^, GMP-certified ET) and placebo (99% microcrystalline cellulose, 1% magnesium stearate) were also provided in coded bottles by Tetrahedron. HPLC grade methanol (A454-4) and acetonitrile (A998-4) were purchased from Fisher Chemical (US). Formic acid (98%; 8841) was purchased from Lancaster (England). All other reagents were purchased from Sigma-Aldrich (US) unless otherwise specified.

### Study design, and ethical approvals

This randomized, double-blinded, placebo-controlled study (ClinicalTrials.gov identifier: NCT03641404) was conducted at the Investigational Medicine Unit, National University Hospital, Singapore in adherence with Good Clinical Practice (GCP). Elderly MCI subjects, 60-90 years of age were randomly assigned to receive either placebo or ET (25mg) capsules (identical capsule appearance, coded by manufacturer), administered orally three times a week, blinded to both subject and administrator for entire duration of the study (Figure 1). Participants visited the study site every 4 weeks (for a total of 14 visits) where they would be provided with sufficient capsules to last until the subsequent visit and underwent safety and compliance monitoring. Blood samples were collected quarterly (visits 1, 4, 7, 10, 14; for assessment of clinical safety assessment and biomarker analyses) and neuro-cognitive assessments were conducted biannually (visits 7 and 14). Baseline cognitive assessments were carried out during volunteer screening. At the trial’s conclusion, subjects underwent a final assessment, followed by a follow-up call one month later to monitor for any delayed adverse effects. The study adhered to ethical standards according to the Declaration of Helsinki and was approved by the Singapore National Healthcare Group, Domain Specific Review Board under protocol number 2017/00982 and Singapore Health Science Authority, Clinical Trial Certificate number CTC1800036.

**Figure 1.**
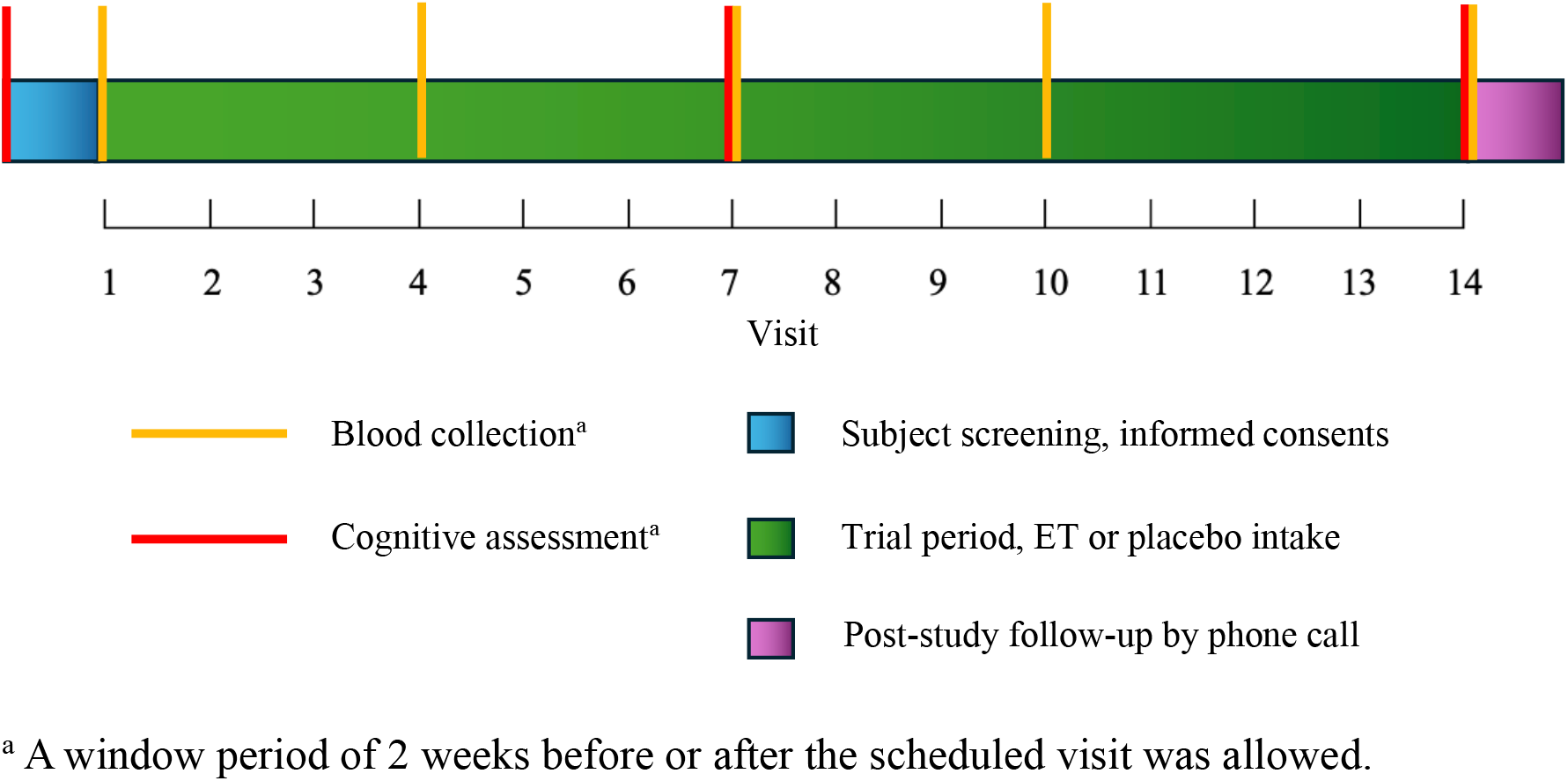
Timeline of the study.

### Subject screening and inclusion/exclusion criteria

Elderly MCI subjects from the Diet and Healthy Ageing study (DaHA)[28], Ageing in a Community Environment Studies (ACES)[29] or Community Health Intergenerational Study (CHI)[30] cohorts (comprised of community-dwelling seniors from the Western regions of Singapore) were contacted (prior consent given to be contacted for future studies) for their potential interest to participate in the study. Given the slow identification rate of MCI subjects, consenting subjects were recruited in staggered enrolment. The recruited volunteers were previously screened (within a 6-month period) using a cognitive assessment battery (comprising the Singapore modified Mini-Mental State Examination; SM-MMSE[31] to assess global cognitive function together with a neurocognitive battery to assess other cognitive domains in detail) and identified as MCI by a panel of 2 senior consultant psychiatrists (EHK, RM) and the cohort PI (LF) according the consensus Petersen’s criteria[32]. General health conditions, including height, weight, blood pressure, age, medical history, dietary intake, were also assessed. Blood and urine samples were collected for biomarker and clinical safety monitoring (blood count, renal and liver function). Inclusion criteria focused on ethnically Chinese individuals aged 60-90 years with MCI. From >1500 subjects screened in the 3 cohorts, 121 MCI subjects meeting the study criteria were contacted to participate. Participants were required to be independent, capable of understanding the study’s demands and of giving informed consent, and free from severe health issues or terminal illnesses. Exclusion criteria included mushroom allergies, history of severe cardiovascular complications, haematological conditions such as anaemia, history of mental or psychiatric illnesses, history of drug or alcohol abuse, concurrent involvement in other studies requiring compound intake in the past 6 months, or any other underlying conditions deemed unsuitable by the investigator. Participants were asked to avoid the use of supplements or Traditional Chinese Medicines and excessive mushroom consumption for the duration of the study, and to report other concurrent medications.

### Study compliance and safety monitoring

Subjects were required to visit the study site every 4 weeks for safety and compliance monitoring. During the study visits, subjects were required to return any unconsumed capsules from the previous batch and provided with a new batch. An administration and adverse event log kept by the subject was checked for evidence of study compliance and safety. In addition to monitoring weight and blood pressure, fasting blood was collected every 3 months to assess safety by monitoring any changes in full blood counts, fasting glucose, and liver and renal function tests. Serum and sodium tubes were sent to the National University Hospital Referral Laboratories (Singapore) for blood parameter analysis including haematological profile, liver panel (albumin, total bilirubin, aspartate transaminase (AST), alanine transaminase (ALT), alkaline phosphatase (ALP)), renal panel (sodium, potassium, urea, creatinine), and fasting glucose. Those failing to adhere to the administration schedule in the first 3 months or demonstrating abnormal blood parameters were withdrawn from the study.

### Blood sampling, preparation, and storage

Fasting venous blood samples was collected in 1x K2-EDTA spray-coated vacuum tube (Becton Dickinson, US), 1x clot activator-treated vacuum tube (Becton Dickinson, US) and 1x sodium fluoride-treated plasma tube (Becton Dickinson, US) for whole blood/plasma, serum, and fasting plasma glucose measurement, respectively. Plasma samples were separated by centrifuging the whole blood samples in EDTA tubes at 1000 x g for 15 min, which were stored at -80°C in 250μl aliquots supplemented with 2μl of 10mM butylated hydroxytoluene (antioxidant) and 1μl of 25mM indomethacin (cyclooxygenase inhibitor) to prevent ex vivo autooxidation.

### Neuro-cognitive assessment

Cognitive functions were assessed during screening, at visit 7 (26 weeks) and at visit 14 (52 weeks) using SM-MMSE modified for Chinese subjects[31], Rey Auditory Verbal Learning Test (RAVLT), Digit Span (WAIS-III UK version), Colour Trials Tests (CTT), Block Design (WAIS-III UK version), Semantic Fluency, Symbol Digit Modality Test (SDMT), Boston Naming Test and Clinical Dementia Rating Scale (CDRS). Z-scores of the test results were calculated using the age- and education-adjusted local norms [33].

### Plasma ET, hercynine, and S-methyl ET measurement by LC-MS/MS

Plasma samples were analysed following previously optimised methods[34]. Briefly, 10μl of plasma was mixed with 100μl methanol containing ET-d9 and hercynine-d9 as internal standards. The sample was mixed and incubated at -20°C for 2h followed by centrifugation. The supernatant was dried under a stream of nitrogen gas, resuspended in 100μl of dH2O, and transferred to a salinized glass insert in a sampling vial (Agilent Technologies, US) for analysis by LC-MS/MS on the Agilent 1290 UPLC and 6460-QQQ MS (Agilent Technologies, US).

### Enzyme-linked immunosorbent assays (ELISA) for plasma biomarkers

ELISA kits for human tumour necrosis factor-α (TNF-α, ab181421), interleukin-18 (IL-18, ab215539), brain-derived neurotrophic factor (BDNF, ab212166), and protein carbonyls (ab238536) were purchased from Abcam. ELISA kits for neurofilament light-chain (NfL, E- EL-H0741) were from Probioscience Technologies. All samples were measured in duplicate, according to the manufacturer’s protocols. All the ELISA assays were colorimetric, with absorbances read at 450nm (BioTek Synergy H1). Concentrations were calculated using appropriate calibration curves.

### Sample size calculations

Our original sample size calculations (conducted by JL) were based on composite cognitive outcomes from a standard neuropsychological assessment battery (from the Finnish Geriatric Intervention Study to Prevent Cognitive Impairment and Disability [35]). Based on these cognitive outcomes, 44 MCI subjects per arm were required for a statistical power of 80% to detect a z-score difference of 0.3 over the one-year intervention, using a conventional alpha of 0.05. Given an estimated attrition rate of 20%, 106 subjects were needed for the study. However, given the complications in recruitment (see Discussion), and failing to hit the intended target, we have chosen to assess this data as a pilot study which demonstrates the safety and potential efficacy of ET.

### Statistical analysis

The statistical analysis adopted an intention-to-treat approach that included data from all participants, including the early withdrawals, to preserve randomness and provide a conservative estimate of the treatment effects. Data are expressed as mean ± standard deviation unless specified, n=11 for the ET-supplemented arm and n=8 for the placebo arm. Paired mixed-effect analysis, which deals with missing data, was used for paired comparison among study time points, followed by Dunnet’s pairwise comparison against baseline with the following annotation: \*\*\**p*<0.001; \*\*\*\**p*<0.0001. Outliers were identified by ROUT method (Q=0.1). Statistical analysis was conducted using GraphPad Prism (Version10).

## Results

### Participant demographic characteristics at baseline

More than 1500 subjects from the three community cohorts were screened, of which 121 identified MCI subjects meeting the study criteria were contacted for interest to participate in the study. However, only 19 were enrolled and 14 completed the study. The poor uptake of the study was in part due to COVID-19 related fears amongst the elderly as the pandemic hit soon after the commencement of the study (also leading to two voluntary withdrawals). Of the five withdrawn, two were due to health complications unrelated to the study and three were due to voluntary withdrawal or non-compliance. Among the 14 completed subjects, some scheduled measurements were missed due to restrictions during the COVID-19 pandemic. The enrolled participants all demonstrated MCI at baseline (as diagnosed by the clinical assessment team) with a median CDR-SOB of 0.5 (*p*=0.001, Wilcoxon signed-rank test against 0) [36]. No significant differences were observed in baseline plasma levels of ET and its metabolites, hercynine and S-methyl ET, between the treatment groups (Table 1). Baseline plasma ET levels demonstrated no significant correlation with mini-mental state examination (MMSE) scores or age (Figure 2a&b). No significant differences were observed between plasma ET levels in males (872.46nM) and females (828.49nM) (Figure 2c). Similarly, levels of the ET metabolites showed no age or gender differences.

**Figure 2.**
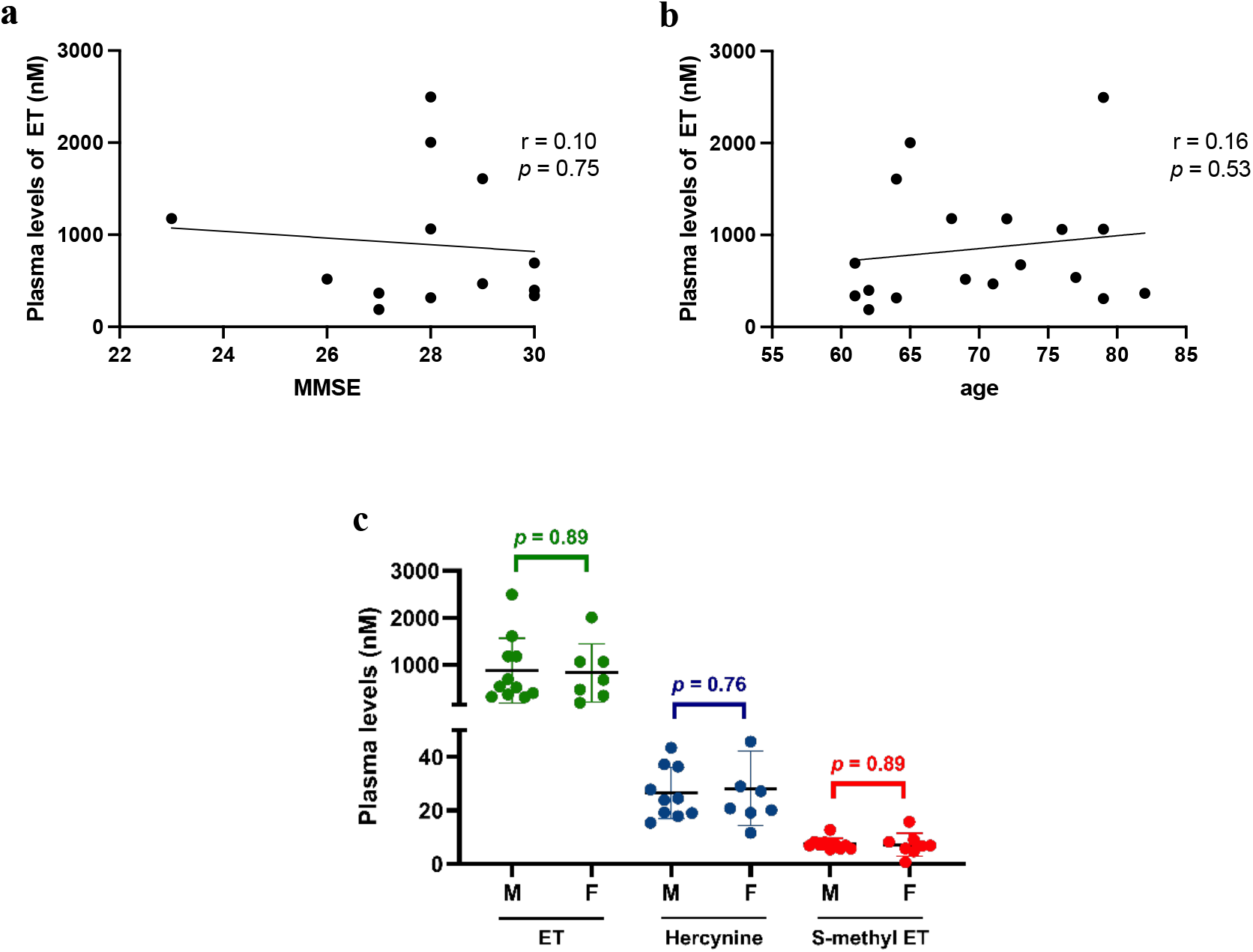
Correlation of ergothioneine with key demographic characteristics at baseline. The correlations of plasma levels of ET with a) MMSE and b) age, showing Pearson’s correlation coefficients (r), trendlines plotted by simple linear regression, and *p* values from statistical comparisons of the slopes against 0. C) Gender differences in baseline plasma levels of ET and its metabolites compared by unpaired student’s t-test (M, male; F, female).

**Table 1.** Baseline characteristics of participants.

|  | Total (n=19) | Study groups |  |  |
| --- | --- | --- | --- | --- |
|  |  | ET (n=11) | Placebo (n=8) | *p* value<sup>a</sup> |
| Age(year) | 69.89±6.97 | 70.55±7.63 | 69.00±6.76 | 0.65 |
| Male, n(%) | 11(58%) | 8(73%) | 3(38%) | - |
| Height(cm) | 162.84±8.19 | 164.91±6.70 | 160.00±9.61 | 0.21 |
| Weight(kg) | 60.63±9.16 | 62.11±7.77 | 58.59±11.02 | 0.42 |
| SBP(mmHg) | 145.37±21.16 | 146.91±23.04 | 143.25±19.61 | 0.72 |
| DBP(mmHg) | 74.00±11.28 | 72.27±10.01 | 76.38±13.16 | 0.45 |
| CDR-SOB, median(IQR) | 0.5(0.625) | 0.5(0.5) | 1(0.5) | 0.093 |
| MMSE | 27.93±1.86 | 27.86±2.67 | 28.00±0.58 | 0.89 |
| Physical activity<sup>b</sup>, median(IQR) | 4(2.25) | 4(2.5) | 4(2.75) | 0.89 |
| Social activity<sup>b</sup>, media (IQR) | 2.5(2) | 2.5(2) | 2.5(1.75) | 0.93 |
| Plasma ET(nM) | 855.36±643.29 | 698.32±415.42 | 1102.13±876.72 | 0.20 |
| Plasma Hercynine(nM) | 27.22±11.34 | 25.10±8.56 | 30.54±14.87 | 0.34 |
| **Plasma S-methyl ET(nM)** | 7.35±3.15 | 7.40±2.10 | 7.27±4.56 | 0.93 |

<sup>a</sup>Student t-test and Mann-Whitney test were used to compare means and medians, respectively, of ET and placebo groups.
<sup>b</sup>Activity frequency scale: 1=never or rarely, 2=more than once a month but less than once a week, 3=one to three times a week, 4=four to six times a week, 5=daily.
Abbreviation: SBP, systolic blood pressure; DBP, diastolic blood pressure; CDR-SOB, Clinical Dementia Rating-Sum of Boxes; IQR, inter-quartile range; MMSE, Mini-Mental State Examination.

### Plasma levels of ET, hercynine, and S-methyl-ET increased following ET intake

At baseline, the average plasma levels of ET, hercynine, and S-methyl-ET of the study population were at 855.36nM, 27.22nM, and 7.35nM, respectively (Figure 3). In subjects with ET supplementation, the ET level increased almost 5-fold to 3.96μM by visit 7 and continued to increase linearly to 6.86μM by visit 14 (Figure 3a). Hercynine and S-methyl-ET, also increased over time, albeit to a smaller extent, and reached 125.15nM and 24.82nM at visit 14, respectively (Figure 3b&c). No significant changes in levels of the three compounds were observed in the placebo group throughout the study.

**Figure 3.**
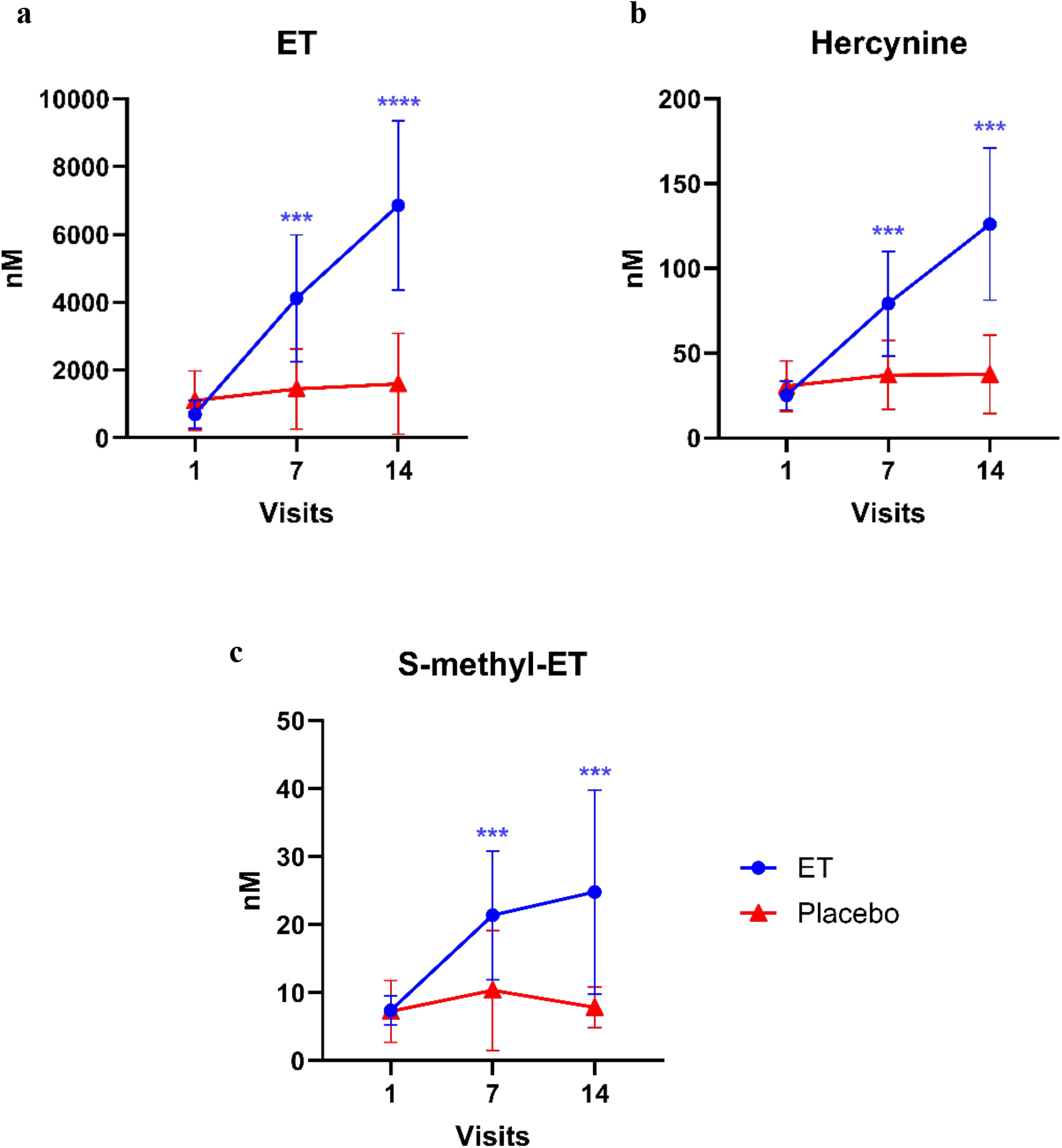
Levels of ET and its metabolites across the study. Plasma levels of a) ET, b) hercynine, and c) S-methyl-ET of each of the study arms at visits 1, 7, and 14. Mixed-effect analysis was used for longitudinal comparison within each of the study arms, followed by Dunnett’s post-hoc test. Asterisks denote significant differences compared to visit 1 (baseline) of the respective study arm \*\*\**p*<0.001, \*\*\*\**p*<0.0001.

### Safety profile of ET

ET administration did not cause significant changes in whole blood parameters, liver function markers, renal function markers, or fasting glucose levels (Table 2). Participants in both ET and placebo groups recorded a lower total white blood cell count compared to baseline at visit 7, both of which recovered subsequently. The reasons for this anomaly are unclear but values were all still within the expected range for their age.

**Table 2.**
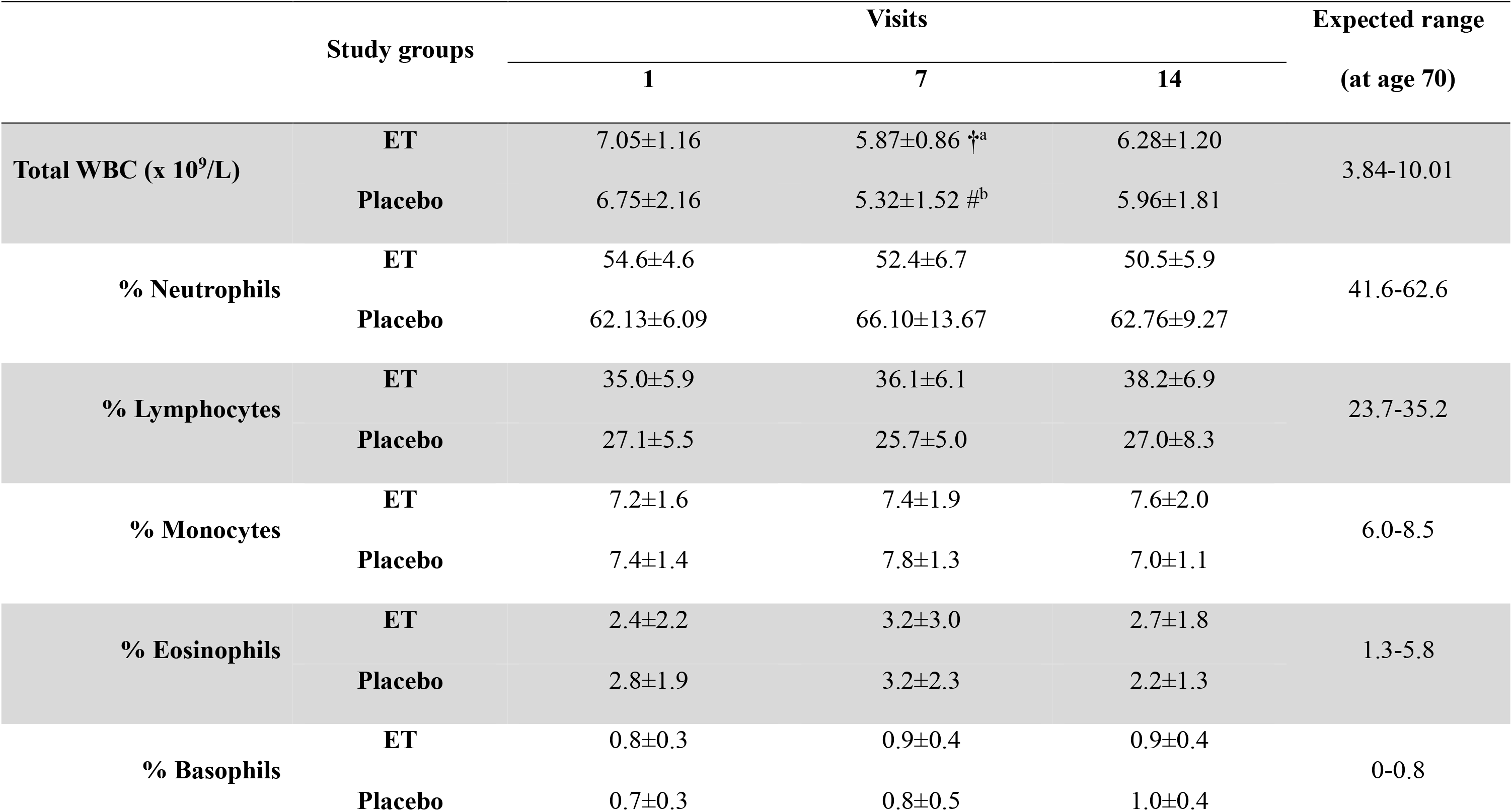
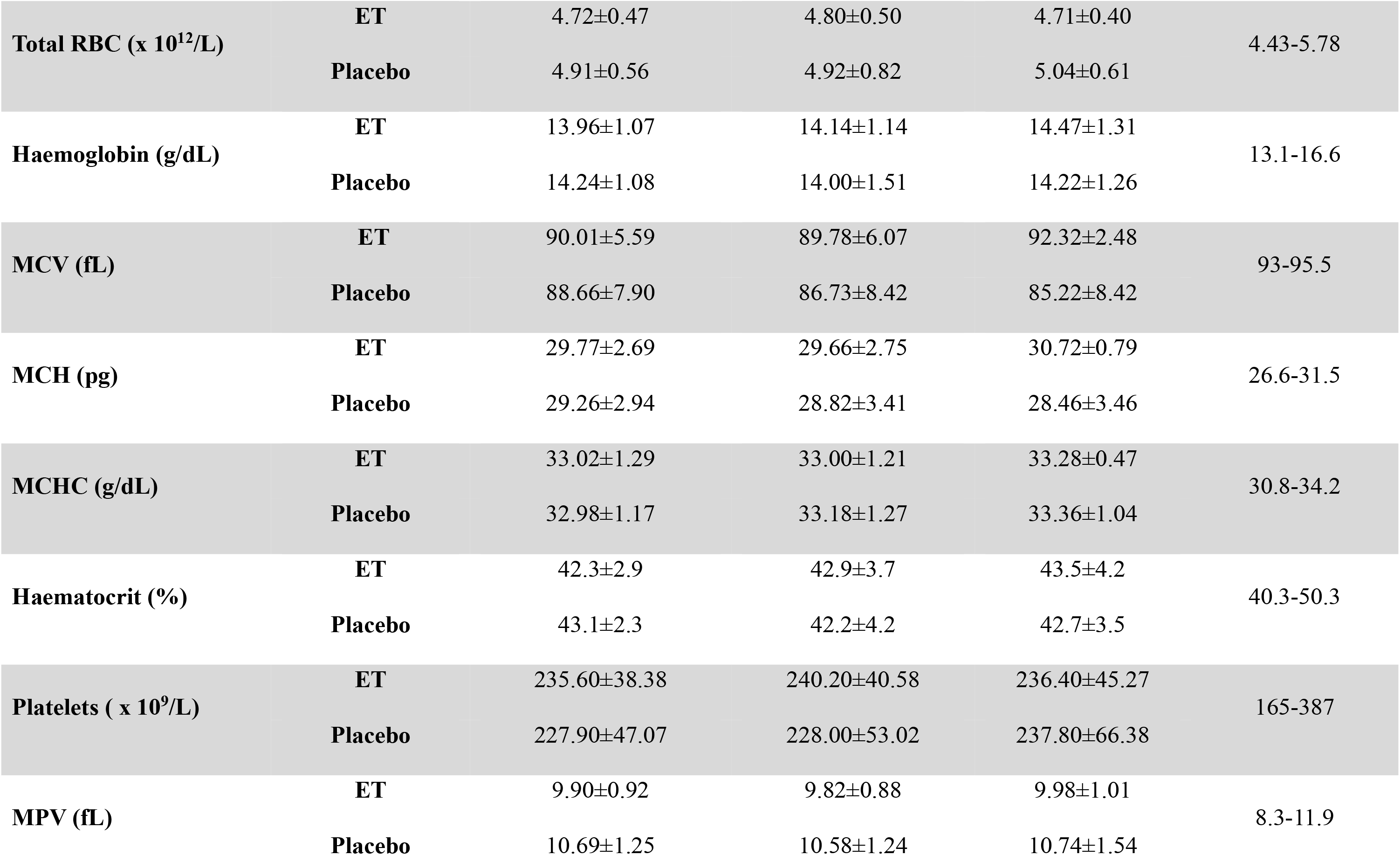
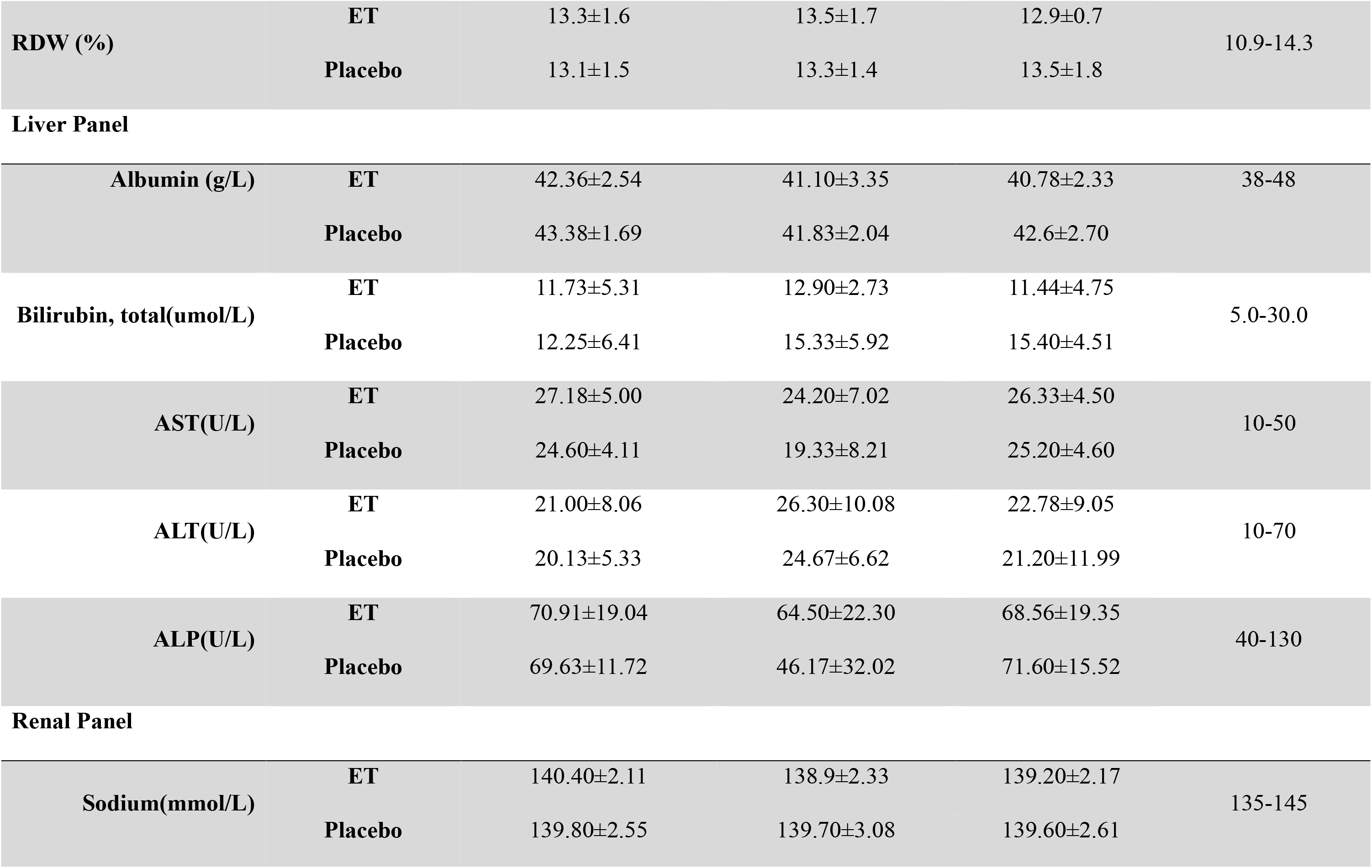
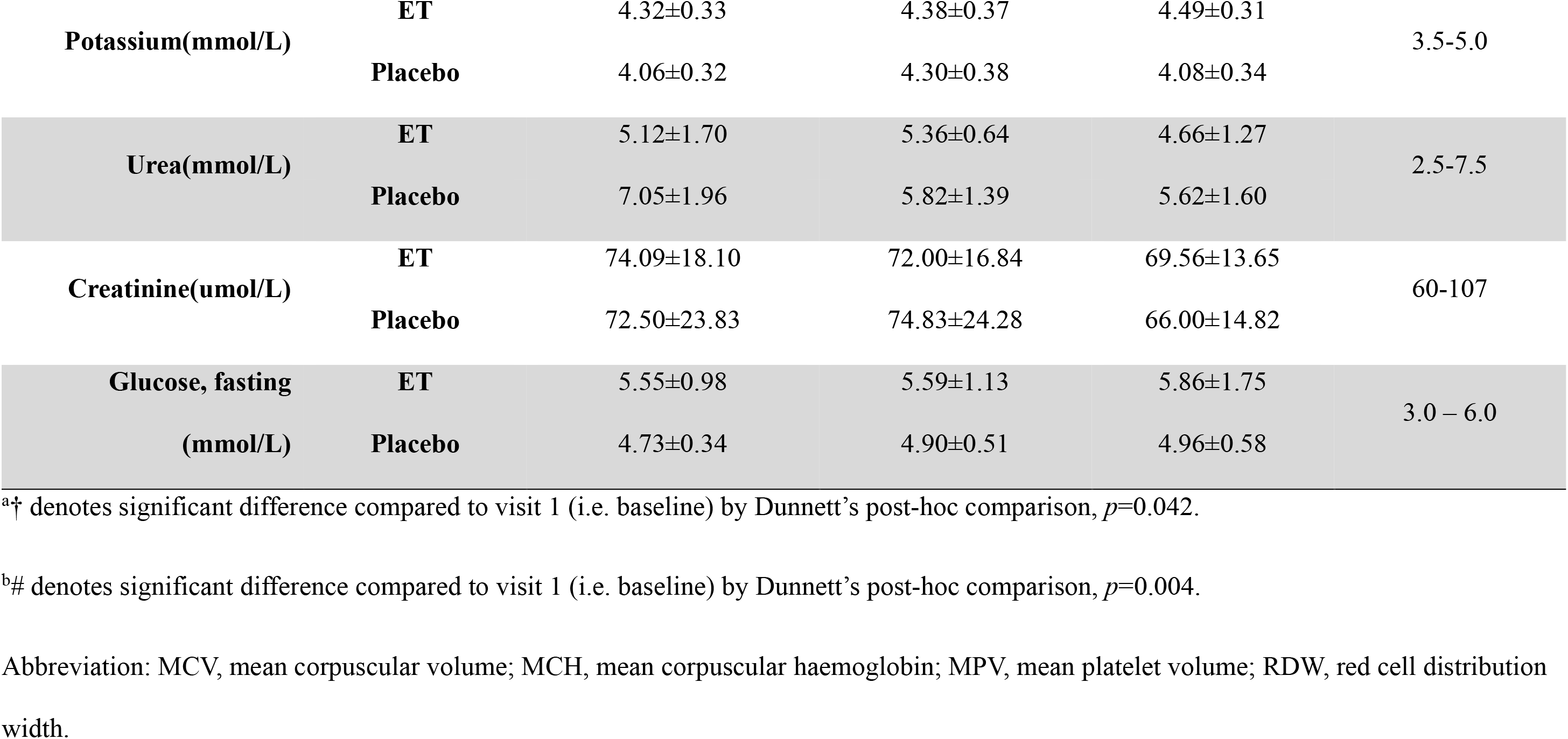
Blood markers for safety profile of prolonged ET consumption

|  | Study groups | Visits |  |  | Expected range<br>(at age 70) |
| --- | --- | --- | --- | --- | --- |
|  |  | 1 | 7 | 14 |  |
| Total WBC (x 10<sup>9</sup>/L) | ET | 7.05±1.16 | 5.87±0.86 †<sup>a</sup> | 6.28±1.20 | 3.84-10.01 |
|  | Placebo | 6.75±2.16 | 5.32±1.52 #<sup>b</sup> | 5.96±1.81 |  |
| % Neutrophils | ET | 54.6±4.6 | 52.4±6.7 | 50.5±5.9 | 41.6-62.6 |
|  | Placebo | 62.13±6.09 | 66.10±13.67 | 62.76±9.27 |  |
| % Lymphocytes | ET | 35.0±5.9 | 36.1±6.1 | 38.2±6.9 | 23.7-35.2 |
|  | Placebo | 27.1±5.5 | 25.7±5.0 | 27.0±8.3 |  |
| % Monocytes | ET | 7.2±1.6 | 7.4±1.9 | 7.6±2.0 | 6.0-8.5 |
|  | Placebo | 7.4±1.4 | 7.8±1.3 | 7.0±1.1 |  |
| % Eosinophils | ET | 2.4±2.2 | 3.2±3.0 | 2.7±1.8 | 1.3-5.8 |
|  | Placebo | 2.8±1.9 | 3.2±2.3 | 2.2±1.3 |  |
| % Basophils | ET | 0.8±0.3 | 0.9±0.4 | 0.9±0.4 | 0-0.8 |
|  | Placebo | 0.7±0.3 | 0.8±0.5 | 1.0±0.4 |  |
| **Total RBC (x 10<sup>12</sup>/L)** | **ET** | 4.72±0.47 | 4.80±0.50 | 4.71±0.40 | 4.43-5.78 |
|  | **Placebo** | 4.91±0.56 | 4.92±0.82 | 5.04±0.61 |  |
| **Haemoglobin (g/dL)** | **ET** | 13.96±1.07 | 14.14±1.14 | 14.47±1.31 | 13.1-16.6 |
|  | **Placebo** | 14.24±1.08 | 14.00±1.51 | 14.22±1.26 |  |
| **MCV (fL)** | **ET** | 90.01±5.59 | 89.78±6.07 | 92.32±2.48 | 93-95.5 |
|  | **Placebo** | 88.66±7.90 | 86.73±8.42 | 85.22±8.42 |  |
| **MCH (pg)** | **ET** | 29.77±2.69 | 29.66±2.75 | 30.72±0.79 | 26.6-31.5 |
|  | **Placebo** | 29.26±2.94 | 28.82±3.41 | 28.46±3.46 |  |
| **MCHC (g/dL)** | **ET** | 33.02±1.29 | 33.00±1.21 | 33.28±0.47 | 30.8-34.2 |
|  | **Placebo** | 32.98±1.17 | 33.18±1.27 | 33.36±1.04 |  |
| **Haematocrit (%)** | **ET** | 42.3±2.9 | 42.9±3.7 | 43.5±4.2 | 40.3-50.3 |
|  | **Placebo** | 43.1±2.3 | 42.2±4.2 | 42.7±3.5 |  |
| **Platelets ( x 10<sup>9</sup>/L)** | **ET** | 235.60±38.38 | 240.20±40.58 | 236.40±45.27 | 165-387 |
|  | **Placebo** | 227.90±47.07 | 228.00±53.02 | 237.80±66.38 |  |
| **MPV (fL)** | **ET** | 9.90±0.92 | 9.82±0.88 | 9.98±1.01 | 8.3-11.9 |
|  | **Placebo** | 10.69±1.25 | 10.58±1.24 | 10.74±1.54 |  |
| **RDW (%)** | **ET** | 13.3±1.6 | 13.5±1.7 | 12.9±0.7 | 10.9-14.3 |
|  | **Placebo** | 13.1±1.5 | 13.3±1.4 | 13.5±1.8 |  |
| Liver Panel |  |  |  |  |  |
| **Albumin (g/L)** | **ET** | 42.36±2.54 | 41.10±3.35 | 40.78±2.33 | 38-48 |
|  | **Placebo** | 43.38±1.69 | 41.83±2.04 | 42.6±2.70 |  |
| **Bilirubin, total(umol/L)** | **ET** | 11.73±5.31 | 12.90±2.73 | 11.44±4.75 | 5.0-30.0 |
|  | **Placebo** | 12.25±6.41 | 15.33±5.92 | 15.40±4.51 |  |
| **AST(U/L)** | **ET** | 27.18±5.00 | 24.20±7.02 | 26.33±4.50 | 10-50 |
|  | **Placebo** | 24.60±4.11 | 19.33±8.21 | 25.20±4.60 |  |
| **ALT(U/L)** | **ET** | 21.00±8.06 | 26.30±10.08 | 22.78±9.05 | 10-70 |
|  | **Placebo** | 20.13±5.33 | 24.67±6.62 | 21.20±11.99 |  |
| **ALP(U/L)** | **ET** | 70.91±19.04 | 64.50±22.30 | 68.56±19.35 | 40-130 |
|  | **Placebo** | 69.63±11.72 | 46.17±32.02 | 71.60±15.52 |  |
| Renal Panel |  |  |  |  |  |
| **Sodium(mmol/L)** | **ET** | 140.40±2.11 | 138.9±2.33 | 139.20±2.17 | 135-145 |
|  | **Placebo** | 139.80±2.55 | 139.70±3.08 | 139.60±2.61 |  |
| **Potassium(mmol/L)** | **ET** | 4.32±0.33 | 4.38±0.37 | 4.49±0.31 | 3.5-5.0 |
|  | **Placebo** | 4.06±0.32 | 4.30±0.38 | 4.08±0.34 |  |
| **Urea(mmol/L)** | **ET** | 5.12±1.70 | 5.36±0.64 | 4.66±1.27 | 2.5-7.5 |
|  | **Placebo** | 7.05±1.96 | 5.82±1.39 | 5.62±1.60 |  |
| **Creatinine(umol/L)** | **ET** | 74.09±18.10 | 72.00±16.84 | 69.56±13.65 | 60-107 |
|  | **Placebo** | 72.50±23.83 | 74.83±24.28 | 66.00±14.82 |  |
| **Glucose, fasting**<br>**(mmol/L)** | **ET** | 5.55±0.98 | 5.59±1.13 | 5.86±1.75 | 3.0 – 6.0 |
|  | **Placebo** | 4.73±0.34 | 4.90±0.51 | 4.96±0.58 |  |

<sup>a†</sup> denotes significant difference compared to visit 1 (i.e. baseline) by Dunnett's post-hoc comparison, $p=0.042$ .
<sup>b#</sup> denotes significant difference compared to visit 1 (i.e. baseline) by Dunnett's post-hoc comparison, $p=0.004$ .
Abbreviation: MCV, mean corpuscular volume; MCH, mean corpuscular haemoglobin; MPV, mean platelet volume; RDW, red cell distribution width.

### ET intake improved performance in certain neuro-cognitive assessments (NCA)

We evaluated the cognitive performance of the subjects using a neuro-cognitive assessment battery consisting of 10 test components, at visit 7 and visit 14 (6 and 12 month, respectively). Baseline scores obtained during the subject screening process (visit 0) within 6 months prior to the commencement of our study, were considered the reference levels. SDMT tests were not conducted during pre-screening. At the baseline, no significant differences in the Z-scores of all NCA components were observed between placebo and ET-supplemented groups. Following ET intake, an increase in Z-scores was observed in RAVLT assessments (immediate and delayed recalls), which evaluates learning ability and memory (Figure 4a&b). In contrast, no significant increase was observed in Z-scores across all NCA components, in the subjects given the placebo. A decrease in the Z-score of CTT1 was seen at visit 7 after ET intake but returned to baseline Z-scores by visit 14 (Figure 4e). Block design scores, which assess visual-motor coordination and problem-solving skills, declined over time in both ET and placebo groups (Figure 4g). Although not measured at baseline, Z-scores of SDMT (written format) in the ET group showed a trend to increase from visit 7 to visit 14 (Figure 4i).

**Figure 4.**
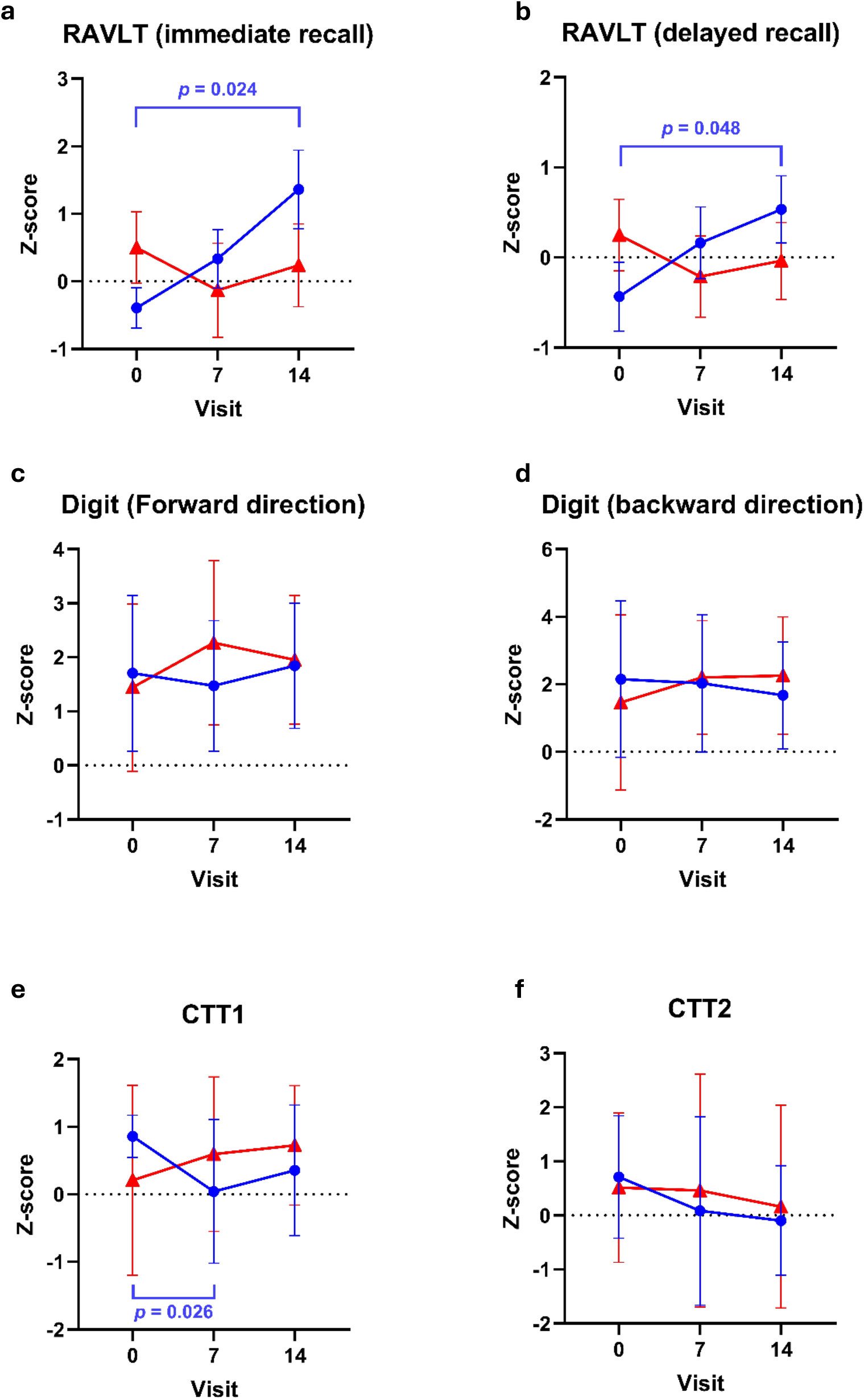
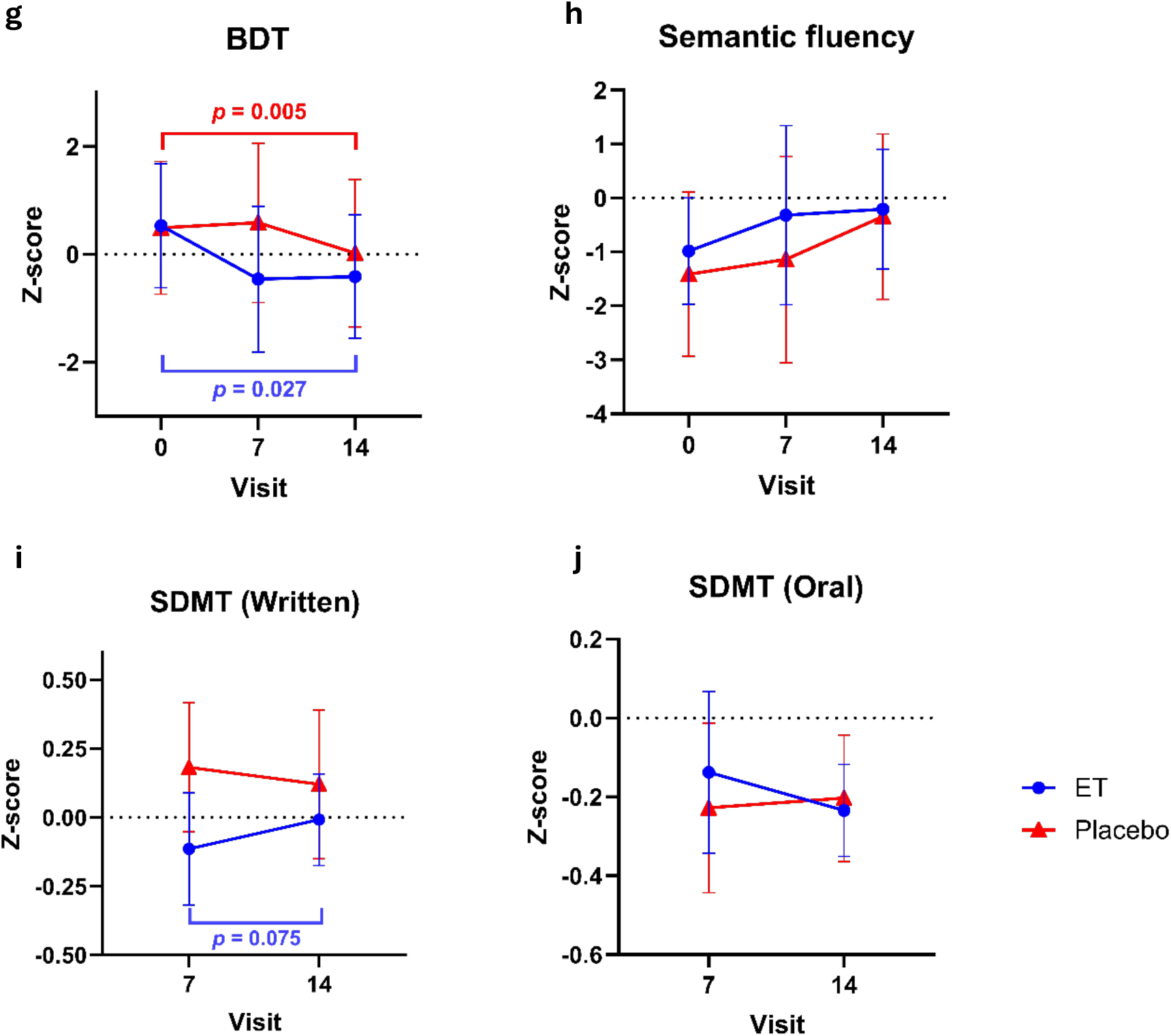
Z-scores of NCA components across the study. Longitudinal data on Z-scores reflecting the performance on a-b) RAVLT, c-d) digit span, e-f) Color trail test, g) block design test (BDT), h) semantic fluency test, and i-j) symbol-digit modality test (SDMT). Mixed-effect analysis was used for longitudinal paired comparison within each of the study arms, followed by Dunnett’s post-hoc test, with only the significant differences (*p*<0.05) annotated.

### ET intake attenuated the plasma level of neurofilament light chain (NfL)

No significant differences in the plasma biomarkers were observed at baseline between ET and placebo arms. Following ET consumption in the MCI subjects, no significant changes in the plasma levels of NfL (a biomarker of neuronal injury) was seen over the 12-month study duration, with a trend to decreasing levels but this was not significant (Figure 5a). Conversely, a significant increase in plasma NfL was observed in the placebo group by visit 14. One data point was identified as an outlier by Prism likely due to technical error and hence the subject was completely removed from the analysis. Both ET and placebo consumption did not affect the plasma levels of BDNF, TNF-α, and IL-18 as markers of neurological function and inflammation, and protein carbonyls, as an index of oxidative protein damage (Figure 5b-e) [37].

**Figure 5.**
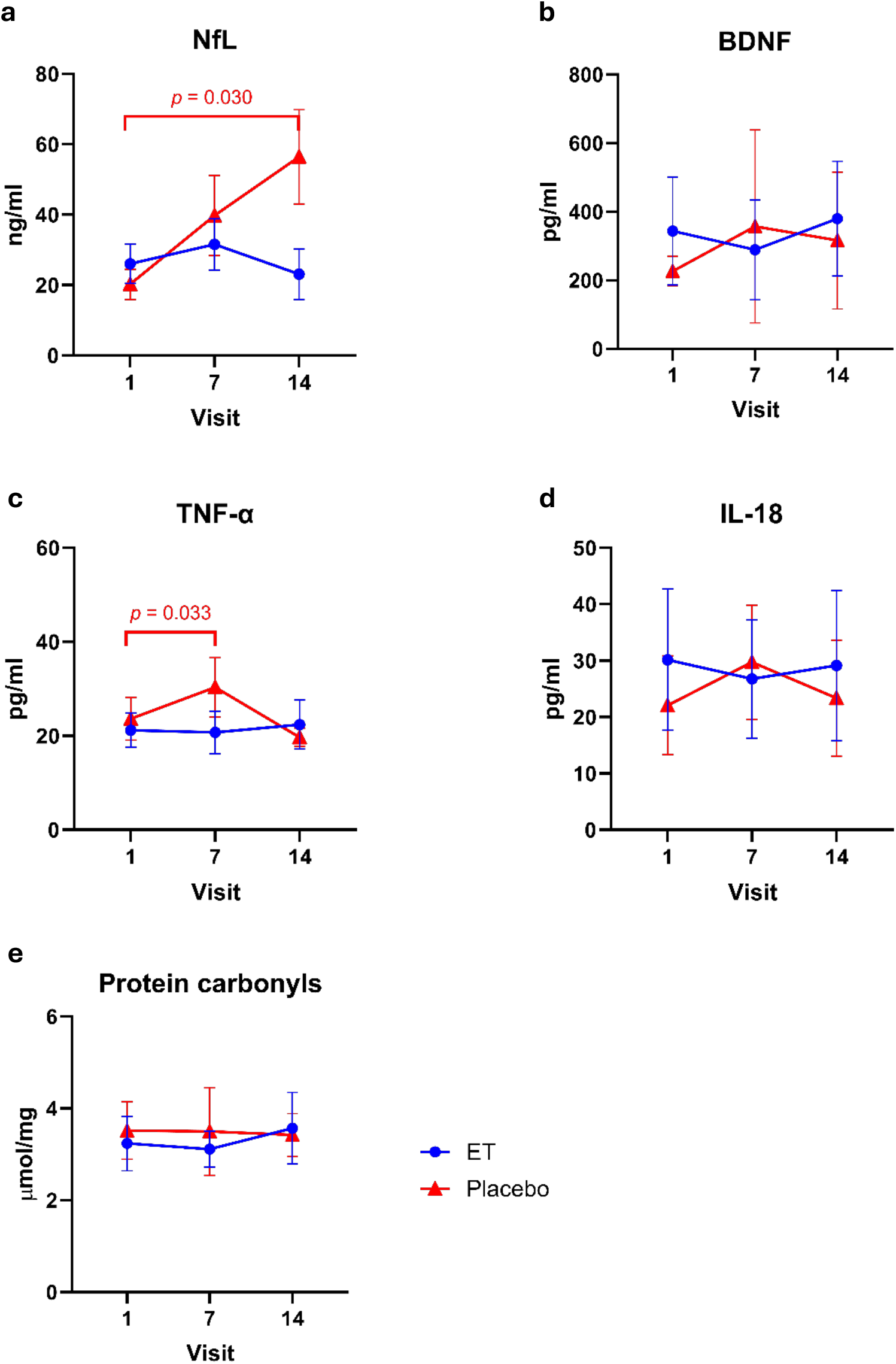
Plasma biomarkers associated with pathological processes of dementia. Longitudinal data on Z-scores reflecting the performance on a-b) RAVLT, c-d) digit span, e-f) Color trail test, g) block design test (BDT), h) semantic fluency test, and i-j) symbol-digit modality test (SDMT). Mixed-effect analysis was used for longitudinal paired comparison within each of the study arms, followed by Dunnett’s post-hoc test, with only the significant differences (*p*<0.05) annotated.

## Discussion

While we have previously demonstrated the association between low plasma levels of ET and individuals with MCI or dementia[23–25], no studies have directly investigated the safety and efficacy of ET supplementation in elderly individuals with MCI. Here we report an interventional pilot study that explores ET’s neuroprotective potential in seniors demonstrating early indications of cognitive impairment. Our findings confirm the strong safety profile of ET, with no observable adverse reactions or changes in clinical parameters over a year-long administration period in elderly individuals.

Plasma levels of ET and its metabolites, namely hercynine and S-methyl ET, were measured at each study time point to verify ET uptake by the subjects. A continuous and significant increase of plasma ET levels was observed with year-long regular (3 times weekly) consumption of ET, increasing linearly with no sign that levels had reached a plateau. These results demonstrate the avid uptake and retention of ET by the body and extend findings from our previous study demonstrating a 3-fold increase in plasma ET levels in healthy human young adults after a 7- day consumption of ET (25mg). The avid uptake and retention of ET is attributed to the expression of the ET transporter, OCTN1, in the human intestinal tract and proximal renal tubules, respectively[15, 38]. We have demonstrated the ability of ET to accumulate in many mouse organs including the brain, following oral administration[39]. The presence of OCTN1 and ET in the central nervous system in laboratory animals and in humans reveals ET’s capability to cross the blood-brain barrier, supporting its potential to be neuroprotective[13].

Concurrently, we evaluated the cognitive performance of our subjects using a comprehensive neuro-cognitive assessment battery. Preliminary outcomes reveal that ET supplementation resulted in a gradual improvement in RAVLT immediate recall and delayed recall testing, which became significantly higher than baseline at visit 14 (Figure 4a&b). However, there was no increase in the Z-scores in the placebo group, indicating the improvements were likely attributable to ET (rather than to practice effect or other extrinsic factors). We also observed a stabilization of plasma levels of NfL (Figure 5a), a biomarker of axonal damage whereby elevated levels are indicative of neuronal function loss and progression of neurological disorders such as dementia[40, 41], in subjects consuming ET. However, mean levels of plasma NfL increased in the placebo group, which was significant at the 12-month timepoint.

The RAVLT recall tests examine the learning ability of adults which involves the formation and consolidation of episodic memory in the medial temporal lobe, including the hippocampus[42]. According to the Hebb’s rule, memory and learning require strengthening and stabilization of pre- and postsynaptic activities associated with the same input and stimulus[43]. Studies in mice suggest that ET promotes hippocampal maturation and improves memory and learning as well increasing synaptic formation in hippocampal cells[44]. Furthermore, early AD features compromised synaptic plasticity and long-term potentiation (LTP) due to the production and accumulation of toxic Aβ oligomers[45, 46]. Indeed, addition of ET to hippocampal slices isolated from APP/PS1 mice, a humanised early AD model, sustained a higher LTP compared to control (reviewed in [13]). Oral administration of ET to various mouse models of AD can also decrease neurodegeneration and memory and learning or cognition deficits [20–22]. The observed improvement in RAVLT and the stabilization of plasma NfL levels with ET-supplementation in MCI subjects suggest that ET may decrease neuronal injury, thus preserving the synaptic plasticity and LTP, therefore better learning abilities and memory. By contrast, the performance on block design task in the placebo group worsened over time, and ET did not mitigate the change. No significant changes were seen in other cognitive domains such as working memory, mental flexibility, executive function, and processing speed.

The plasma markers of oxidative protein damage and inflammation did not show significant differences compared to the placebo group, consistent with our previous study showing only small effects of ET on oxidative damage markers in plasma and urine in healthy subjects[34]. Of course, peripheral levels of these biomarkers may not reflect events happening in the brain.

Besides monitoring of adverse events, the safety of ET administration was also monitored through quarterly assessment of various clinical parameters including liver and renal function and haematological assessment. No differences were observed in both placebo and ET administration group relative to baseline, and no adverse reactions from ET administration were recorded, consistent with previous data that ET is safe for prolonged supplementation. There was one anomaly whereby a reduction in total white blood cell count was observed in both the ET and placebo groups at the mid-point of the study, both of which returned to baseline values thereafter. However, as this occurred in both study arms, it is likely not due to ET.

Despite our promising findings, consistent with the proposed neuroprotective actions of ET[13], our study is subjected to several limitations, the most severe is the small number of participants, precluding generation of conclusive data as originally planned. The COVID-19 pandemic greatly impacted the study including the screening and enrolment of MCI subjects, hampered our data collection as some participants missed scheduled assessments due to COVID-19- related restrictions and reluctance of elderly subjects to attend the clinic located near a hospital, two voluntary withdrawals at the onset of the pandemic and a very significantly decreased willingness to participate in the study in screened MCI subjects (>85% rejection by screened subjects). Consequently, the small size and composition of our cohort should be carefully considered when interpreting our work. Future studies will require a larger and more diverse study population and longer study duration to validate our findings, but our study lays the background for these and reinforces the safety of ET.

In conclusion, this pilot study confirms the safety and possible efficacy of ET and lays the groundwork for future large-scale trials that assess its role in cognitive health and as a therapeutic for neurodegenerative disorders. ET emerges not only as a potential preventive agent for cognitive decline but also as a therapeutic agent conferring direct neuroprotection in MCI patients. Further research should aim to decipher the mechanisms underpinning ET’s cognitive benefits, particularly its functional implications in AD patients when taken together with first-line disease-modifying therapies.

## Data Availability

All data produced in the present study are available upon reasonable request to the authors.

## Acknowledgements

The authors wish to thank Jean Claude Yadan (ERGOLD; formally Tetrahedron, Paris, France) for the kind provision of the encapsulated ET (25mg, Ergoneine®) and placebo used in this study as well as analytical standards for ET and related metabolites.

